# Hypertension and Renin–Angiotensin–Aldosterone System Inhibitors in Patients with Covid-19

**DOI:** 10.1101/2020.04.24.20077388

**Authors:** Andrew Ip, Kaushal Parikh, Joseph E. Parrillo, Shivam Mathura, Eric Hansen, Ihor S. Sawczuk, Stuart L. Goldberg

**Affiliations:** Division of Outcomes and Value Research, John Theurer Cancer Center at Hackensack University Medical Center, Hackensack, NJ, USA; COTA, Boston, MA, USA; Hackensack University Medical Center, Hackensack, NJ, USA; Hackensack Meridian School of Medicine at Seton Hall University, Nutley, NJ, USA; Hackensack Meridian Health, Edison, NJ, USA

## Abstract

**Introduction:** COVID-19 disproportionately affects those with comorbidities and the elderly. Hypertension is the most common pre-existing condition amongst COVID-19 patients. Upregulation of the renin-angiotensin-aldosterone system (RAAS) is common in hypertensive patients and may promote inflammation and ensuing cytokine storm in COVID-19. It is unknown whether RAAS inhibition with ACE1 inhibitors or angiotensin-receptor blockers (ARB) can be harmful or beneficial.

**Methods:** Within Hackensack Meridian Health network, the largest healthcare provider in New Jersey, we performed a retrospective, multicenter, convenience sampling study of hospitalized COVID-19 patients. Demographics, clinical characteristics, treatments, and outcomes were manually abstracted. Fishers exact tests, and logistic regression were performed.

**Results:** Among 3017 hospitalized COVID-19 patients, 1584 (52.5%) carried a diagnosis of hypertension. In the discharged or deceased cohort, the overall mortality was significantly increased at 35% vs 13% among COVID-19 patients with hypertension. However, when adjusted for age, the effect of hypertension on mortality was greatly diminished, with a reduction in odds-ratio by over half; and completely disappeared when adjusted for other major covariates. The mortality rates were lower for hypertensive patients prescribed ACE1 (27%, p=0.001) or ARBs (33%, p=0.12) compared to other anti-hypertensive agents (39%) in the unadjusted analyses. RAAS inhibitor therapy appeared protective compared to other anti-hypertensive agents (p=0.001).

**Conclusions:** While our results are limited by the retrospective nature of our study and by potential confounders, our data argue against a harmful effect of RAAS inhibition and support the HFSA/AHA/ACC joint statement recommending continuing ACE1 and ARB therapy in hypertensive COVID-19 patients.

## INTRODUCTION

Elderly patients, especially with comorbid conditions, have been disproportionately affected by SARS-CoV-2, the causative agent in COVID-19. Hypertension is the most common pre-existing condition among COVID-19 patients and early reports note an association between hypertension and increased mortality.^1^ Upregulation of the renin-angiotensin-aldosterone system (RAAS) occurs in hypertensive patients and may promote inflammation and ensuing cytokine storm in COVID-19.^2^ The spike glycoprotein of SARS-CoV-2 gains entry into cells by binding to angiotensin converting enzyme 2 (ACE2) on epithelial cells of the lungs, intestines and kidneys. Subsequent downregulation of surface ACE2 results in accumulation of angiotensin II (ANGII) with effects on vasoconstriction, inflammation and fibrosis.^2,3^ Pharmacologic inhibition of RAAS with ACE1 inhibitors and angiotensin-receptor blockers (ARBs) increase ACE2 expression and could be harmful or beneficial.^4^ Given these concerns, physicians may be tempted to alter anti-hypertensive therapy during COVID-19, although abrupt withdrawal of RAAS inhibitors has been associated with cardiac instability in patients with cardiomyopathies.^5^

## METHODS

We performed a retrospective, multicenter, convenience sampling study of patients hospitalized with confirmed SARS-CoV-2 within the Hackensack Meridian Health network, the largest healthcare provider in New Jersey. Demographics, clinical characteristics, treatments, and outcomes were manually abstracted from electronic health records. Institutional review board approval was obtained for the database, but subject informed consent was waived as analysis was conducted on secondary use de-identified data. Fishers exact tests, and logistic regression were performed.

## RESULTS

Among 3017 hospitalized COVID-19 patients, 1584 (52.5%) carried a pre-existing diagnosis of hypertension; 794 (50.1%) have been discharged, 422 (26.6%) died from complications, and 368 (23.2%) remain hospitalized. In the cohorts with a known outcome (discharge or death) the overall mortality appeared significantly increased at 35% (422/1216) for individuals with hypertension compared to 13% (140/1112) in COVID-19 patients without hypertension (p=0.0001). However, hypertension was more frequently observed in older populations. When adjusted for age, the effect of hypertension on mortality was diminished (table 1), with a reduction in odds-ratio [unadjusted OR 3.7, 95% CI (2.99-4.58) vs. adjusted OR 1.6, 95% CI (1.23-1.99)]. When additional factors (gender, ICU entry on day of admission, serum ferritin, insulin-dependent diabetes mellitus and cardiac arrhythmia history) are entered into a Cox proportional hazard model the effect of hypertension disappears. ACE1 inhibitors and ARBs were used in 22.8% and 18.0% of hypertensive patients with known outcomes. Treatment by RAAS inhibitors was not associated with detrimental effects and may be protective. The mortality rates were lower for hypertensive patients prescribed ACE1 (27%, p=0.001) or ARBs (33%, p=0.12) compared to other anti-hypertensive agents (39%) in the unadjusted analyses. RAAS inhibitor therapy appeared protective compared to other anti-hypertensive agents (p=0.001) (Table 2).

**Table 1.** Hypertension (HTN) vs. Age Among Patients with Known Disposition (Expired or Discharged)

| Age | HTN<br>Expired | HTN<br>Discharged | No HTN<br>Expired | No HTN<br>Discharged | *p-value |
| --- | --- | --- | --- | --- | --- |
| <= 50 | 6 (5.9%) | 95 (94.1%) | 16 (3.2%) | 491 (96.8%) | 0.236 |
| 51-60 | 28 (14.5%) | 165 (85.5%) | 21 (8.5%) | 225 (91.5%) | 0.066 |
| 61-70 | 83 (27%) | 224 (73%) | 33 (18.4%) | 146 (81.6%) | 0.036 |
| 71-80 | 115 (39.4%) | 177 (60.6%) | 32 (29.9%) | 75 (70.1%) | 0.101 |
| >= 81 | 190 (58.8%) | 133 (41.2%) | 38 (52.1%) | 35 (47.9%) | 0.297 |

\*P-value by Fisher's test, using Bonferroni correction for multiple hypothesis testing to control for false discovery rate, the alpha is set to 0.01 (a p-value would need to be < 0.01 to be statistically significant)

**Table 2.** ACE inhibitor or ARB vs. Other Anti-hypertensives in Hypertension patients.

| Disposition | Total | RAAS inhibition<br>(ACE-i/ARB)* | Other anti-HTN* | p-value** |
| --- | --- | --- | --- | --- |
| Discharged | 730 | 323 (70%) | 407 (61%) | 0.001 |
| Expired | 399 | 137 (30%) | 262 (39%) |  |

\*RAAS = Renin-Angiotensin-Aldosterone System; ACEi = Angiotensin Converting Enzyme 1 inhibitor; ARB = Angiotensin Receptor Blocker; HTN = Hypertension
\*\*P-value by Fisher's exact test

## DISCUSSION

The Heart Failure Society of America (HFSA), American Heart Association (AHA) and American College of Cardiology (ACC) released a joint statement recommending continued use of ACE1 and ARB inhibitors in hypertensive patients with COVID-19.^6^ Our retrospective real-world review of hospitalized patients found that hypertension did not worsen mortality when corrected for age and other key factors. We did not identify harmful effects of RAAS inhibition and there was a suggestion of possible benefit. There is evidence that hypertensive COVID-19 patients have higher ANGII levels contributing to a pro-inflammatory state.^4^ Therefore, ACE1s and ARBs, which decrease ANGII production or antagonize its action by blocking its receptor, might suppress the inflammatory state of COVID-19.^4^ A Chinese retrospective report also noted lower in-hospital mortality with RAAS inhibitors compared to other anti-hypertensive agents.^3^ While our results are limited by the retrospective nature of our study, our data argue against a harmful effect of RAAS inhibition and support the HFSA/AHA/ACC joint statement recommending the continuation of ACE1 and ARB therapy in hypertensive COVID-19 patients.

## Data Availability

This is a institution owned data set. All data can only be shared externally with a data use agreement.

## ACKNOWLEDGEMENTS

The authors wish to thank the nurses, data managers, and physicians who after caring for their patients assisted in the abstraction of the clinical data.

## No Competing interests

All authors have completed the ICMJE uniform disclosure form at www.icmje.org/coi_disclosure.pdf and declare: no support from any organization for the submitted work; no financial relationships with any organizations that might have an interest in the submitted work in the previous three years; no other relationships or activities that could appear to have influenced the submitted work.

